# Plasma cell-free DNA methylome profiling in pre- and post-surgery oral cavity squamous cell carcinoma

**DOI:** 10.1101/2022.08.02.22278241

**Authors:** Krupal B Patel, Tapan A Padhya, Jinyong Huang, Liang Wang, Xuefeng Wang

## Abstract

**Purpose:** Head and neck squamous cell carcinoma cancer (HNSCC), a highly heterogeneous disease that involves multiple anatomic sites, is a leading cause of cancer-related mortality worldwide. Although the utility of noninvasive biomarkers based on circulating cell-free DNA (cfDNA) methylation profiling has been widely recognized, limited studies have been reported so far regarding the dynamics of cfDNA methylome in oral cavity squamous cell carcinoma (OCSCC). It is hypothesized in this study that comparison of methylation profiles in pre- and post-surgery plasma samples will reveal OCSCC-specific prognostic and diagnostic biomarkers.

**Materials and methods:** Matched plasma samples from eight patients with OCSCC were collected at Moffitt Cancer Center before and after surgical resection. Plasma-derived cfDNA was analyzed by cfMBD-seq, which is a high-sensitive methylation profiling assay. Differential methylation analysis was then performed based on the matched samples profiled. As a strategy to further prioritize tumor-specific targets, top differential methylated regions (DMRs) were called by reanalyzing methylation data from paired tumor and normal tissue collected in the TCGA head and neck cancer cohort.

**Results:** In the top 200 HNSCC-specific DMRs detected based on the TCGA dataset, a total of 23 regions reached significance in the plasma-based DMR test. The top five validated DMR regions (ranked by the significance in the plasma study) are located in the promoter regions of genes *PENK, NXPH1, ZIK1, TBXT* and *CDO1*, respectively. The genome-wide cfDNA DMR analysis further highlighted candidate biomarkers located in genes *SFRP4, SOX1, IRF4* and *PCDH17*. The prognostic relevance of candidate genes was confirmed by survival analysis using the TCGA data.

**Conclusion:** This study supports the utility of cfDNA-based methylome profiling as a promising noninvasive biomarker source for OCSCC and HNSCC.

## INTRODUCTION

Head and neck squamous cell carcinoma cancer (HNSCC) is a highly heterogeneous disease that involves multiple anatomic sites including oral cavity, larynx, and pharynx. Outcomes of patients with HNSCC have not improved significantly over the past decade, with an overall five-year survival rate around 50%. There is a pressing need in the area to develop reliable prognostic and diagnostic biomarkers to enable better patient management, from early detection of disease to efficient monitoring of cancer recurrence following treatment. Despite their established role in cancer development^1^, epigenetic biomarkers and DNA methylation (DNAme) remain understudied in HNSCC research. Currently, TCGA-HNSC is the only publicly available resource for mining DNAme patterns in head and neck cancer. Cell free DNA (cfDNA) includes both genetic and epigenetic information^2^ and offers several advantages including monitoring tumor burden^3^, and novel discovery of biomarkers for diagnosis and prognosis.^4^ cfDNA is thought to potentially incorporate metastatic sites thus addressing tumor heterogeneity.^5,6^ Aberrant DNA methylation changes are thought to occur early during tumorigenesis and enables tumor progression^7^ and thus may be a more specific and sensitive approach to identify minimal residual disease and prognosis.^8,9^ While genetic analysis of cfDNA can be challenging due to its low yield and being highly fragmented,^10^ plasma cfDNA next generation assays are starting to be utilized in routine clinical use for solid malignancies such as lung^11^ and colon cancers^12^ to make treatment decisions.

cfDNA has been reported to decrease to background level following surgery.^13^ Therefore, we hypothesized that comparing methylation profiles in pre- and post-surgery plasma samples will help validate HNSCC-specific prognostic and diagnostic biomarkers, and provides an opportunity for novel biomarker discovery. Here, we focus on single anatomic site and assess the feasibility of detecting cfDNA methylome in patients with locoregional oral cavity squamous cell carcinomas (OCSCC) and the methylome dynamics in post-operative setting. A high-sensitive cfDNA methylome profiling technique called cfMBD-seq was applied on collected plasma samples. Different from bisulfite conversion-based sequencing methods, cfMBD-seq capture and quantify methylated DNA by methyl-CpG binding protein (MBD). cfMBD-seq is able to generate high-quality sequencing read with ultra-low amount of input DNA (2-10 ng per ml), and has demonstrated better performance in terms of enrichment of CpG islands compared to similar protocols such as cfMeDIP-seq.^14^ To facilitate cfDNA methylation biomarker prioritization with limited sample size, we first conducted a bioinformatics analysis to detect differentially methylated regions (DMRs) based on the matched tumor-normal tissue data collected from the TCGA-HNSC project. We hypothesized that the top cancer-specific DMRs detected in the TCGA-HNSC cohort will also exhibit differential methylation patterns between before- and after-surgery plasma samples. As an alternative strategy, we performed a genome-wide search of top DMRs based on the matched cfDNA methylation profiles. DMR analyses were conducted on different patient subgroups as a sensitivity analysis considering the presence of patient and sample heterogeneity. Once we identified top DMRs, we also examined their prognostic relevance in the TCGA patient data, as well as their performance in discriminating pre- and post-treatment plasma samples.

## MATERIALS AND METHODS

### Patient sample collection

To detect HNSC-specific cfDNA biomarkers, we studied the plasma samples collected from a cohort of head and neck cancer patients treated at Moffitt Cancer Center (Tampa, USA). In this pilot study, a total of 16 matched plasma samples were collected from 8 patients before and at least 4 weeks after surgery. The basic clinical characteristics of these patients are displayed in **Table 1**. The study was approved by Institutional Review Board at Moffitt. All patients were consented to the protocol and all samples are de-identified during the methylation profiling process and in the downstream analysis.

**Table 1.**
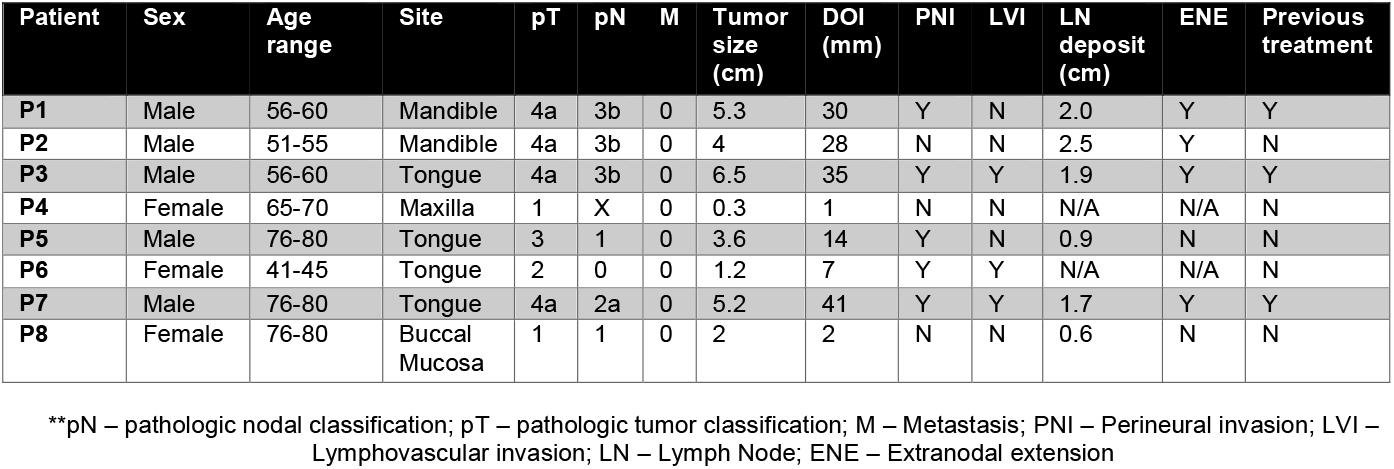
Baseline clinical characteristics and treatment of patients included in the plasma cfMBD-seq analysis.

### TCGA-HNSC analysis

To facilitate cfDNA methylation biomarker discovery, we first conducted a bioinformatics analysis to detect differentially methylated regions (DMRs) based on the tumor tissue methylation data collected from the TCGA-HNSC project. A total of 580 samples were profiled by the Illumina Infinium HumanMethylation450 BeadChip (450K array) in this cohort. Because the goal is to identify cancer-specific regions, our DMR analysis focuses on the 100 paired tumor and normal tissue methylation data collected from 50 patients. We downloaded level 3 DNA methylation data (beta value) from Broad Firehose web portal (gdac.broadinstitute.org) and all clinical data from GDC data portal (portal.gdc.cancer.gov). DMR analysis was performed using the bumphunter function implemented in the R package “minfi”, with the effect size cutoff set at 0.3 and the resampling number at 1000. We restricted downstream analyses on regions > 5 bp in length, in which all regions contain at least two probes (L≥2). The detected regions were annotated against genome build UCSC hg19 by using the annotateDMRInfo function implemented in the “methyAnalysis” package.

### Plasma cfDNA methylome profiling by cfMBD-seq

Cell-free DNA in plasma were profiled by cfMBD-seq, which is an enrichment-based ultra-low input cfDNA methylation profiling method recently developed by our team at Moffitt^14^. Briefly, Maxwell RSC ccf DNA Plasma kit was used to extract the cfDNA from 1 ml of plasma. If one sample contains less than 5ng DNA, we extract DNA from another 1 ml plasma sample. We then combined the cfDNA from the first and second extraction (if needed for a patient sample) for the methylation enrichment and sequencing library preparation. Methylated DNA fragments were enriched and captured by using a MethylCap Kit. cfMBD libraries were prepared and quantified following the steps as previously described in the cfMBD-seq protocol, and sequenced by Illumina NextSeq 500/550 High Output Kit (75 cycles).

After fastq file merging and adapter trimming steps, sequence reads were aligned to hg19 assembly using BWA MEM (v0.7.10). Mapped reads were further sorted and filtered to remove low-quality and duplicated reads using samtools (v1.9) and picard tools (v1.82). R package “MEDIPS” was used to conduct coverage saturation analysis and downstream QC analysis. The “qsea” R package was used to calculate normalized methylation (beta) levels both at a genome-wide level and in targeted ROI regions (such as promoter regions). We applied fitNBglm function in “qsea” to perform the differential coverage analysis. The function fits a negative-binomial model for each genomic widow similar to the differential expression analysis function implemented in the package “edgeR”. To maximize the statistical power, the design matrix in the DMR analysis was formed in a paired DMR setting, in which an additive model formula is formed to include both treatment effect and subject effect. For statistical testing, a reduced model was fit by the function without the treatment term and the p-value is generated by comparing the likelihood ratio of the models against a Chi-square distribution.^15,16^ To remove potential noise regions with low coverage, we only consider regions (1kb window) with a minimum of 50 reads in all the DMR tests.

### Prioritizing candidate cfDNA methylation biomarkers

Because the current study has limited sample size and HNSC samples exhibit high heterogeneity in nature, we propose two schemes to efficiently prioritize the most robust cfDNA biomarker panels while minimizing the false discovery markers or ones with weak clinical significance. As illustrated in the Fig 1A, the first biomarker discovery scheme only investigates DMRs that have been detected based on the analysis using the TCGA data. We hypothesize that the top cancer-specific DMRs detected based on the matched tumor-normal tissues will also exhibit differential methylation patterns between before- and after-surgery plasma samples. The stringent genome-wide multiple-testing correction is not required in this setting because it becomes a targeted biomarker validation analysis. In the second scheme, we perform genome-wide differential coverage analysis on cfDNA methylation data by only considering regions that are located in or nearby the promoter of known genes (defined as 5kb upstream and 2kb downstream of the TSS regions). Furthermore, as will be explained more in the next section, we considered different patient subgroups for the pre- and post-treatment DMR analysis. In each test, regions with an adjusted p-value less than 0.1 or unadjusted p-values less than 1×10-6 will be reported. Similar to the TCGA analysis, the detected regions were annotated using the “methyAnalysis” package. In summary, we reason that both schemes are useful in identifying promising targets that could be further tested as diagnostic and prognostic markers in managing HNSC patients.

**Figure 1.**
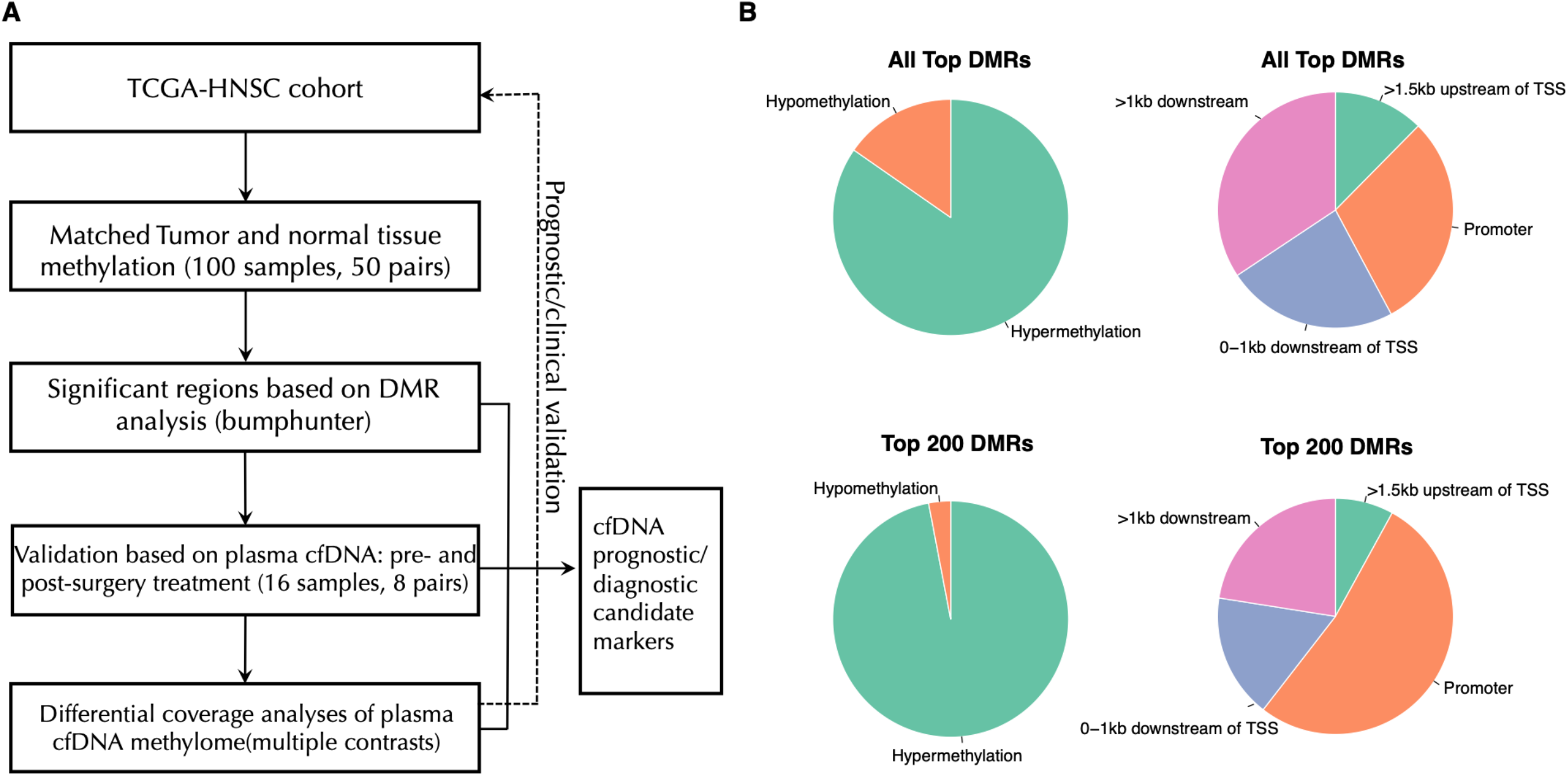
Overview of the proposed DMR analysis on OCSCC plasma samples. A. The experimental design and overall analytical workflow for cfDNA methylation profiling on pre- and post-treatment OCSCC patient samples. B. Pie charts showing the distribution of methylation status and genomic locations in the top detected DMRs.

## RESULTS

### Candidate regions based on TCGA-HSNC analysis

The DMR analysis by comparing the TCGA-HNSC matched tumor and normal methylation profiles identified a total of 1468 significant regions (effect size cutoff of 0.3 and FWER adjusted p value<0.01) (Supplementary Table 1). As shown in Figure 1B, the majority (84.7%) of these top regions are hypermethylated DMRs; and more than half of regions are located in promoter (29.8%) or nearly (0-1kb) downstream regions of TSS (23.5%). When we narrow the list to the top 200 DMRs only, the proportion of hypermethylated DMRs and DMRs in promoter region further increased to 97% and 52.5%, respectively. The average length of top 200 DMRs is 461 bp. The top five DMRs in the promoter region are located in genes *MARCHF11, ZNF154, ELMO1, ADCYAP1* and *PIEZO2*. A summary of top DMRs and their associated genes is provided in Table 2. Interestingly, we observed that many zinc-finger genes were enriched in the top DMR list, to only list those in the top 100 list: *ZNF154, ZNF582, ZNF135, ZNF136, ZNF577, ZNF781, ZNF529, ZNF132, ZNF85, ZNF583, ZNF471*, and *ZNF665*.

**Table 2.**
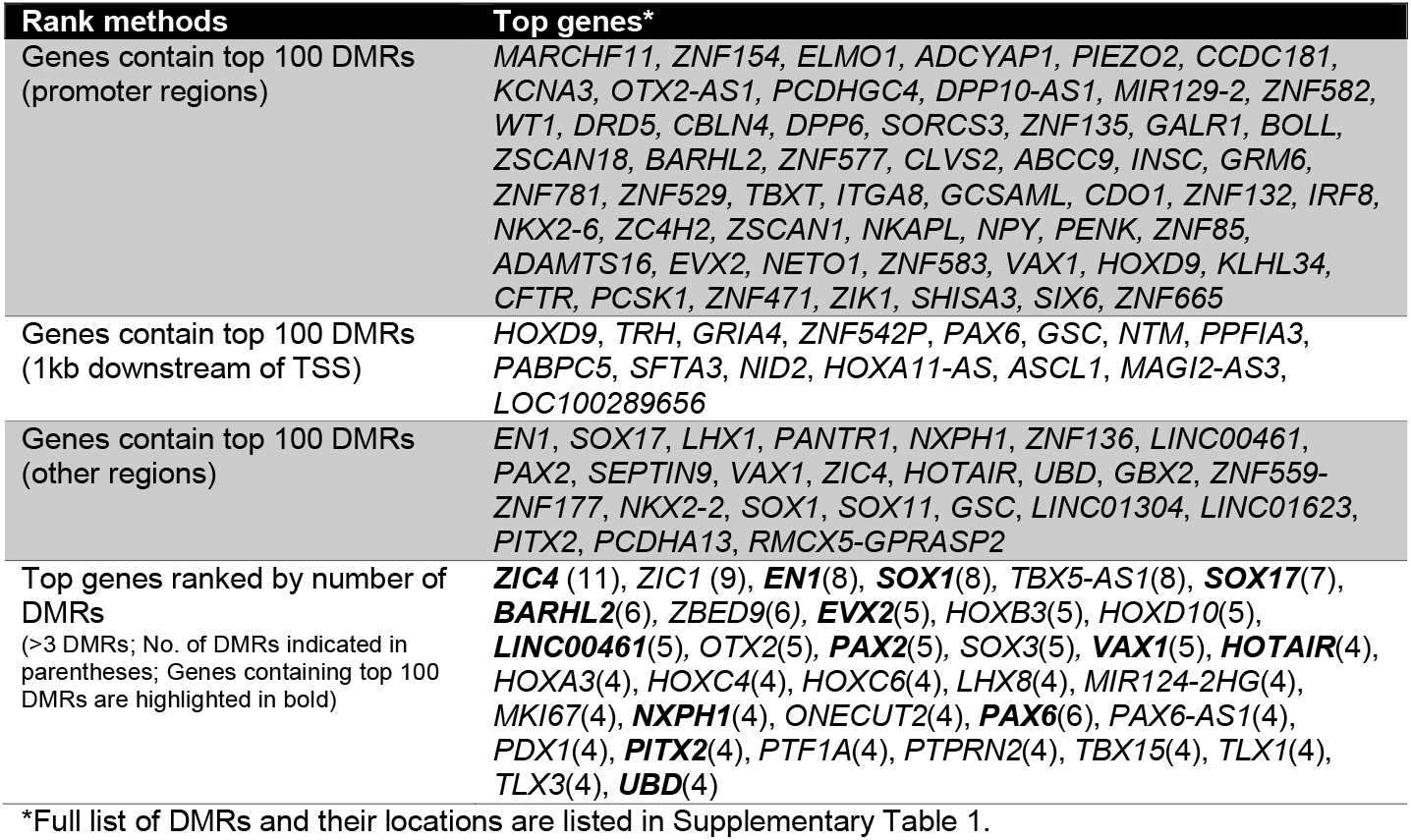
Table of top genes containing significant DMRs based on TCGA-HNSC matched tumor and normal tissue methylation profiles.

### Targeted DMR validation on plasma cfDNA methylomes

In this section, we focus on validating the significant DMRs detected between tumor and normal tissues. We first performed a paired DMR screening test by comparing the matched pre- and post-treatment cfMBD-seq data, using a fixed window size at 1kb. In the top 200 DMRs detected in the TCGA dataset, we found a total of 23 overlapping gene regions reached significance (p<0.05) in the plasma DMR test (Supplementary Table 2), including the two regions in *ZNF154* and *ELMO1* (top five candidate DMRs from the TCGA analysis). Another two regions *ADCYAP1* and *PIEZO2* from the top five candidate DMRs also reached marginal significance (p-value ∼0.06). The normalized methylation levels at these top regions across the 16 plasma samples are depicted in Figure 2 (ranked by plasma DMR p-values). The top five validated regions are located in the promoter regions of genes *PENK, NXPH1, ZIK1, TBXT* and *CDO1*. A clear pattern revealed by Figure 2 is that the TCGA-based DMRs showed the best discriminating power between pre- and post-treatment samples for patients P1 and P7, followed by P4 and P8. Overall, patients P1 and P7 showed the most drastic methylation changes between the pre- and post-treatment samples across most targeted DMRs, potentially due to the fact that both patients (both are male) and had T4 tumors. The pattern of methylation is heterogenous between different patients as evidenced by results in Figure 2. For example, changes in methylation in patient P4 are more pronounced in DMRs in *NXPH1, ZIK1* and *CNTNAP2*, while changes in P2 (also with a T4 tumor) are more pronounced in *HOXD9, TMEM132C* and *SORCS3*.

**Figure 2.**
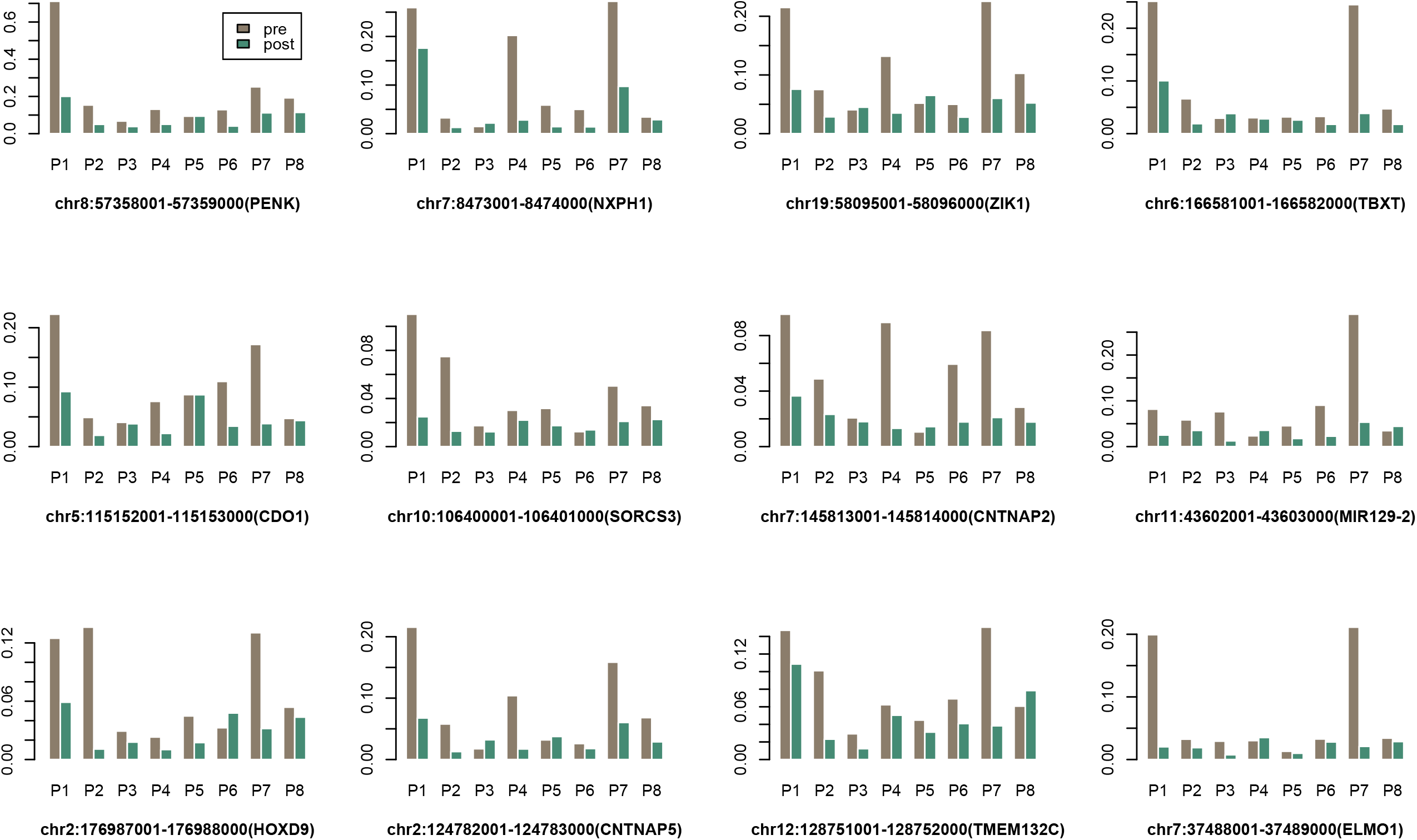
The normalized methylation levels of top DMRs across the matched plasma samples from 8 patients.

### Genome-wide cfDNA DMR analysis on patient subgroups

Given the heterogeneity of head and neck cancer at the molecular level and the limited sample size, we reason that it is more powerful to perform DMR analyses based on subsets of patients based on their clinical characteristics. The patient subset information and the resulted top significant or suggestive DMRs (defined as a p-value < 0.1 level for adjusted p-value or at 10^−7^ level for unadjusted p-value) are listed in Table 3. It is interesting to note that the DMR test based on the four patients (P1, P4, P7 and P8) that had the most concordant patterns with TCGA data resulted in the highest number of significant DMRs, followed by the two patient subsets containing T4 tumors (P1, P2, P3 and P7). We excluded patient P5 in most tests (except for the tongue-site subgroup) because the genome-wide PCA analysis (Supplementary Figure S1) indicated that the pre-treatment cfDNA methylation profiles could be a potential outlier, which may explain why there was no significant DMR when all patients are included in the test. The multiple-contrast DMR tests can also be considered as sensitivity analyses that provide further confidence for overlapped findings. It was observed that the genomic regions in gene *PENK, SFRP4* and *SOX17* were selected in at least three tests, suggesting they might be further prioritized for biomarker validation. Figure 3A shows the detailed methylation levels in top regions that were identified based on the TCGA-concordant subgroup, including two top-ranked validated DMRs listed in Figure 2 (*PENK* and *ZIK1*). Similar to the pattern observed in Figure 2, patients P1 and P7 showed the most drastic changes between the pre- and post-treatment samples. Through assessing the gene expression levels of these top genes in TCGA-HNSC data, we observed that four genes (*ZIK1, IRF4, PCDH17* and *PENK*) demonstrated significant or suggestive association with patient overall survival, as shown in Figure 3B-E. Collectively, these findings suggest that these four genes may have tumor suppressor functions and are often hypermethylated in HNSC tumor samples or pre-treatment plasma samples.

**Table 3.** Table of top cfDNA DMRs identified based on comparing pre- and post-treatment cfDNA methylation profiles by considering different patient subgroups.

| Patient group/subgroup | Subset rationale | Top cfDNA DMRs (hg19) <sup>†</sup> |
| --- | --- | --- |
| Paired samples from P1, P4, P7, and P8 | Patterns most consistent with TCGA results | chr7:37956001-37957000 (*SFRP4*)<br>chr8:67873001-67874000 (*TCF24*)<br>chr2:45171001-45172000 (*SIX3* *)<br>chr11:32458001-32459000 (*WT1-AS*)<br>chr8:53478001-53479000 (*ALKAL1*)<br>chr8:57358001-57359000 (*PENK*)<br>chr13:93879001-93880000 (*GPC6*)<br>chr8:55370001-55371000 (*SOX17*)<br>chr4:183062001-183063000 (*TENM3-AS1* *)<br>chr1:119535001-119536000 (*TBX15* *)<br>chr19:58095001-58096000 (*ZIK1*) |
| Paired samples from P1, P2, P3, and P7 | Patients with T4 tumors (male patients) | chr7:37956001-37957000 (*SFRP4*)<br>chr1:47696001-47697000 (*TAL1*)<br>chr5:1594001-1595000 (*SDHAP3*)<br>chr7:87257001-87258000 (*ABCB1/RUNDC3B*)<br>chr8:57358001-57359000 (*PENK*) |
| Paired samples from P3, P5, P6, and P7 | Tongue site only | chr11:43602001-43603000 (*MIR129-2*) |
| P4, P6, and P8 | Female patients only | No region significant at $10^{-7}$ level |
| Paired samples from P1, P2, P3, P4 and P7 | All T4 patients+ P4 (signature patient) | chr7:37956001-37957000 (*SFRP4*)<br>chr8:57358001-57359000 (*PENK*)<br>chr8:55370001-55371000 (*SOX17*)<br>chr3:129693001-129694000 (*TRH*)<br>chr14:48143001-48144000 (*MDGA2* *)<br>chr14:48145001-48146000 (*MDGA2*) |
| Paired samples from P1, P2, P3, P4, P6, P7, and P8 | All patients except P5 (potential outlier) | chr8:57358001-57359000 (*PENK*)<br>chr7:37956001-37957000 (*SFRP4*)<br>chr8:55370001-55371000 (*SOX17*) |

<sup>†</sup>Unless otherwise specific, DMRs are significant at 0.1 level for adjusted p-value or at $10^{-7}$ level for unadjusted p-value. DMRs not in the promoter region of the gene are indicated by “\*”.

**Figure 3.**
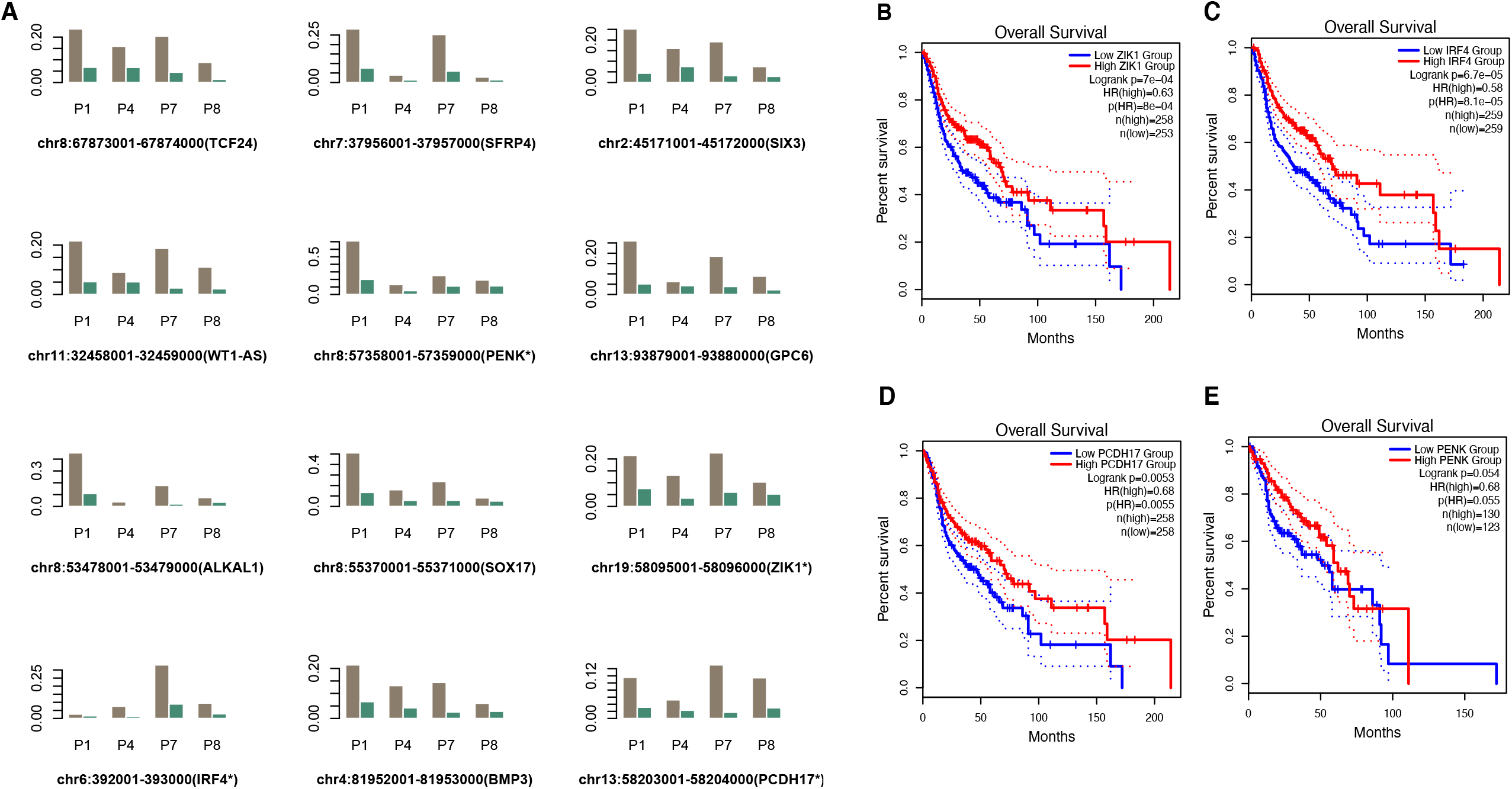
Prioritizing cfDNA DMRs based on the four-patient subgroup. A. Methylation levels in top regions that were identified based on the TCGA-concordant patient subgroup (P1, P4, P7 and P8) B. (C,D,E) Kaplan-Meier plots validating the prognostic significance of four genes that include detected DMRs (based on the gene expression and survival data from the TCGA-HNSC data).

### Clustering analysis of targeted plasma cfDNA methylation regions

Finally, we tested the performance of top DMRs (as well as the model saturation in terms of the number of biomarkers included) in discriminating pre- and post-treatment plasma samples. The two heatmaps in Figure 4 illustrate the unsupervised clustering results generated based on top 30 DMRs and top 200 DMRs (from the DMR test using all samples but P5), respectively. It shows that the top 30 regions (most of them are hypermethylated in pre-treatment samples) are already sufficient to separate pre- and post-treatment plasma samples except for the P5 pre-treatment sample. This was expected, because the global PCA analysis also indicated that this sample could be a potential outlier. But when top 200 regions were included, this sample, together with all other samples, can be correctly separated.

**Figure 4.**
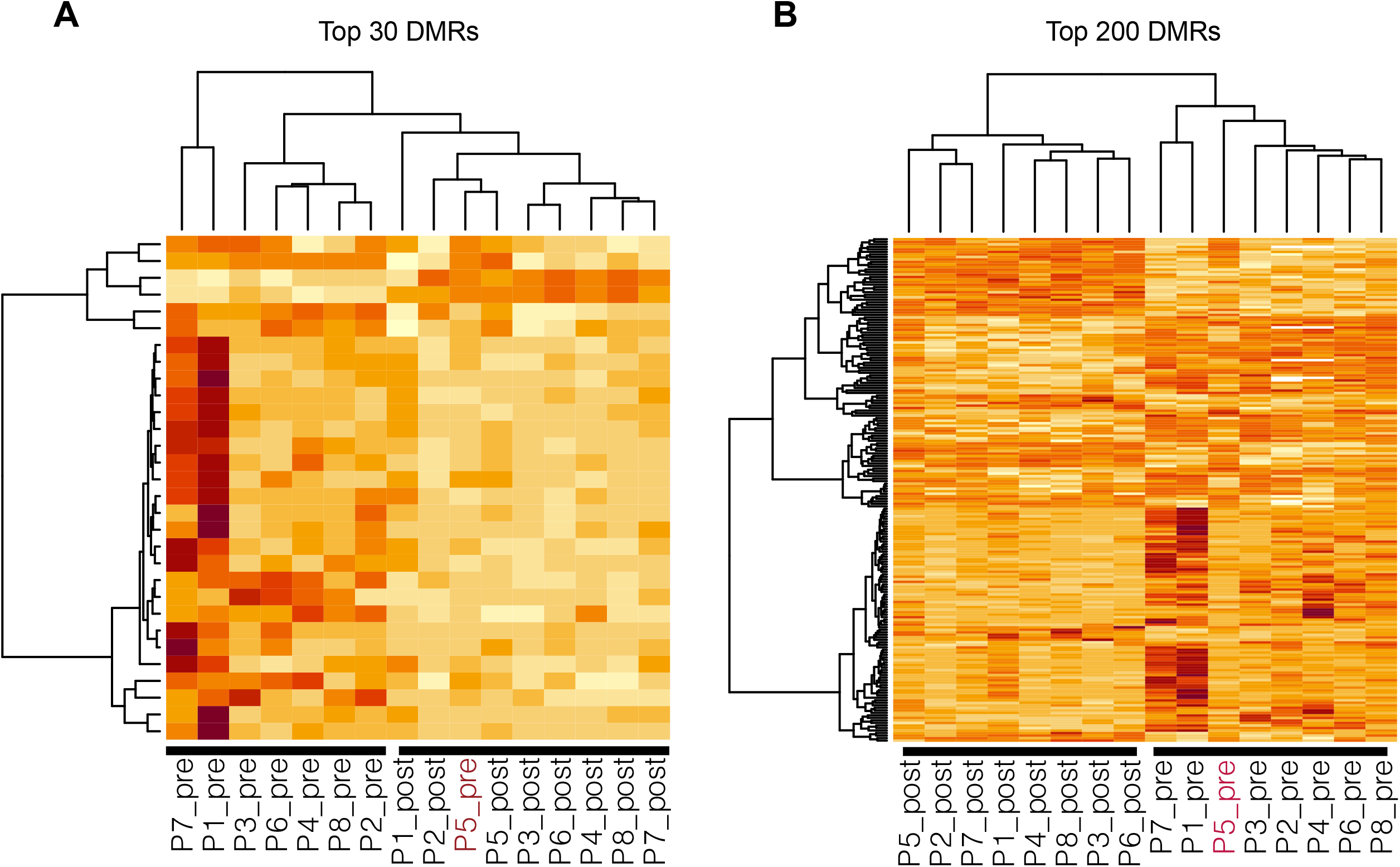
The unsupervised clustering results generated based on top 30 DMRs (A) and top 200 DMRs (B) showing the separation of pre- and post-treatment samples.

## DISCUSSION

While the current study does not have sufficient sample size to comment on prognostic value of cfDNA methylation, in this study, we successfully demonstrate the feasibility of isolating cfDNA from plasma and a two-pronged approach in identifying top candidate biomarkers by first identifying DMRs from the TCGA dataset and then validating them in our cfDNA samples.

A biomarker for minimal residual disease is advantageous, especially in the setting of oral cavity squamous cell carcinoma patients, a population where 5-year survival rates are estimated between 40-60% for patients with advanced-stage disease, and the majority of the recurrence occurs in the first 2 years.^17–20^ Moreover, if the biomarkers can predict tumor immune response it would be even more preferable.^21^ DNAme has the potential to incorporate both of the above features in addition to it being highly tissue specific,^22^ and more responsive to genetic variations and environmental exposure.^23,24^ HNSCC is a heterogenous disease and thus focusing on cfDNA DNAme may be advantageous to increase specificity. Previous studies have focused on targeted panels of methylation in either serum/plasma of HNSCC patients.^25–31^ Recently, a study by Burgener et al. did demonstrate tumor-naïve detection of ctDNA by simultaneously profiling mutations and methylation.^32^

In our study, we identified hypermethylation of zinc finger genes, such as *ZNF154*, in the TCGA tumor samples compared to normal tissue. This is consistent with previous studies where their expression has been shows to correlate with tumor suppressor activity.^33^ *ZNF154* has been reported as diagnostic marker in liquid biopsies in multiple cancers.^34^

Our study also suggests that top 30 DMRs are sufficient to differentiate between pre-treatment and post-treatment samples suggesting that a signature based on these 30 DMRs maybe sufficient to determine minimal residual disease. Many genes in the top DMR list have also been suggested as liquid biopsy methylation biomarkers in other cancer types, such as ZNF154 fir multiple cancers,^34^ *ELMO1* for gastric cancer^35^, suggesting that they are reliable cancer-relevant epigenetic biomarkers. The promoter methylation level of *IRF4* and *PCDH17* have been previously reported as potential liquid biopsy biomarkers for colorectal cancer^36,37^ and bladder cancer.^38^

The top five validated regions were located in the promoter regions of genes *PENK, NXPH1, ZIK1, TBXT* and *CDO1*. Through our analysis 4 candidate genes were identified that may have prognostic value in addition to their role in determining minimal residual disease – *ZIK1, IRF4, PCDH17* and *PENK*. ZIK1 (ZNF762) is part of the Zinc Finger protein group with a KRAB-A domain and found on chromosome 19.^39^ KRAB box-A is a transcription repressor module^39^ and it is plausible that ZIK1 is epigenetically regulated tumor suppressor gene.^33,40,41^ Interferon regulatory factor 4 (IRF4) is a member of the Interferon family and is specifically expressed in lymphocytes^42^ regulating immune responses, immune cell proliferation and differentiation.^43^ While its role in hematologic malignancies has been described previously,^44^ *IRF4* expression in lung adenocarcinoma has been associated with favorable prognosis.^45^ Protocadherin 17 (PCDH17) is part of the cadherin superfamily responsible for cell adhesion and possible tumor growth, migration and invasion.^46,47^ PCDH17 methylation has been noted in urological cancers including esophageal,^48^ gastric,^49^ colon,^49^ and bladder cancers.^50^ Proenkephalin (PENK) is expressed in nervous and neuroendocrine systems as part of the opioid pathway^51^, but is also involved in cell cycle regulation and implicated in head and neck,^52^ gastric,^53^ colon,^54^ breast,^55^ pancreatic,^56^ osteosarcoma,^57^ and bladder^58^ cancers. NXPH1 is primarily expressed in nervous system and is a secreted glycoprotein that forms complexes with alpha neurexins – a group of protein that promote adhesion between dendrites and axons^59^. In breast cancer samples, NXPH1 methylation levels were lower compared to normal tissues and was more likely to be methylated in low-grade dysplasia than in high-grade dysplasia.^60^ In prostate cancers with Gleason score ≥ 7, NXPH1 expression level was upregulated and was incorporated in a 10 gene signature that predicted biochemical recurrence.^61^ However, a negative correlation was noted in patients with pancreatic cancer with regards to lymph node metastasis.^62^ NXPH1 methylation has also been implicated in neuroblastoma^63^ and was incorporated in a 5 gene prognostic signature where it was down regulated suggestive of playing a tumor suppressive role.^64^ This suggests that tissue specific changes maybe at play. T-box transcription factor T (TBXT) expression is implicated in mesodermal specification during vertebrate development^65^ and is epigenetically silenced in human fetus development at 12 weeks.^66^ TBXT expression has been reported in a number of solid malignancies including head and neck^67^, lung,^68^ breast,^69^ colon,^69^ prostate^70^ and chordoma^71^ – with hypothesis that it promotes epithelial-mesenchymal transition and targeting it may help in cancer control.^72^ Promoter methylation *CDO1* has also been previously identified as diagnostic biomarkers in lung cancer.^9^

Limitations of this study include limited samples size to draw definitive prognostic conclusion. cfDNA has been correlated with overall stage and subsite. Our study included primarily advanced stage disease and mainly oral cavity squamous cell carcinomas. It is well established that advanced stage cancers^9^ and different subsites will have different methylation pattern^41^ and lymphatic drainage, thus it is plausible that similar results may not be evident in lower stage disease. In summary, we identified multiple candidate DMRs that allowed distinction between pre-treatment and post-treatment samples suggesting its utility for minimal residual disease and potential as prognostic biomarker.

## Data Availability

All data produced in the present study are available upon reasonable request to the authors.

## Notes

**Funding support:** This work has been supported in part by Moffitt Clinical Science Fund; a National Institute of Health grant R01DE030493; and Moffitt’s Biostatistics and Bioinformatics Shared Resources, Tissue Core, and Genomics Core Facilities at the H. Lee Moffitt Cancer Center & Research Institute, an NCI-designated Comprehensive Cancer Center (P30-CA076292).

**Conflicts of interest disclosure:** None declared

### Competing Interest Statement

The authors have declared no competing interest.

### Funding Statement

This work has been supported in part by Moffitt Clinical Science Fund; a National Institute of Health grant R01DE030493; and Moffitt s Biostatistics and Bioinformatics Shared Resources, Tissue Core, and Genomics Core Facilities at the H. Lee Moffitt Cancer Center & Research Institute, an NCI-designated Comprehensive Cancer Center (P30-CA076292).

### Author Declarations

The study was approved by Institutional Review Board at Moffitt. All patients were consented to the protocol and all samples are de-identified during the methylation profiling process and in the downstream analysis.

