## Supplementary Figure S1 for "Plasma cell-free DNA methylome profiling in pre- and post-surgery oral cavity squamous cell carcinoma"

**Supplementary Figures**

**A**

**B**
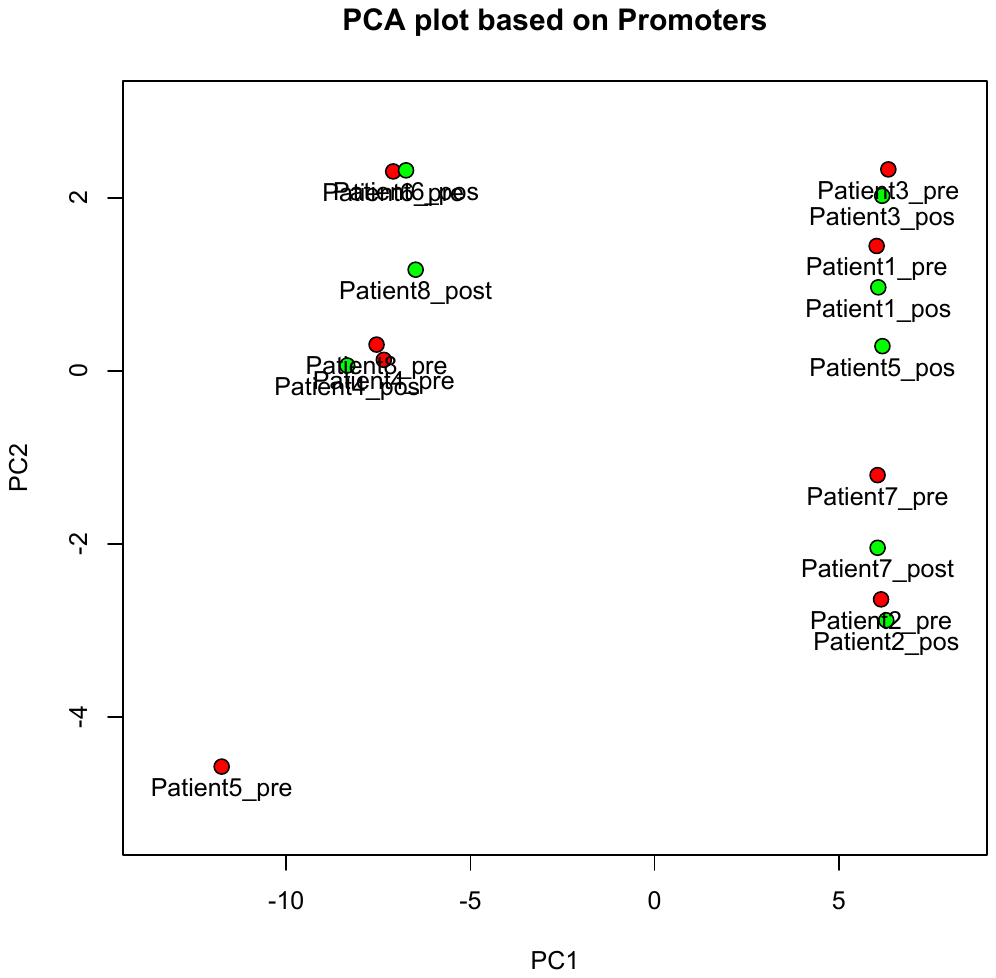

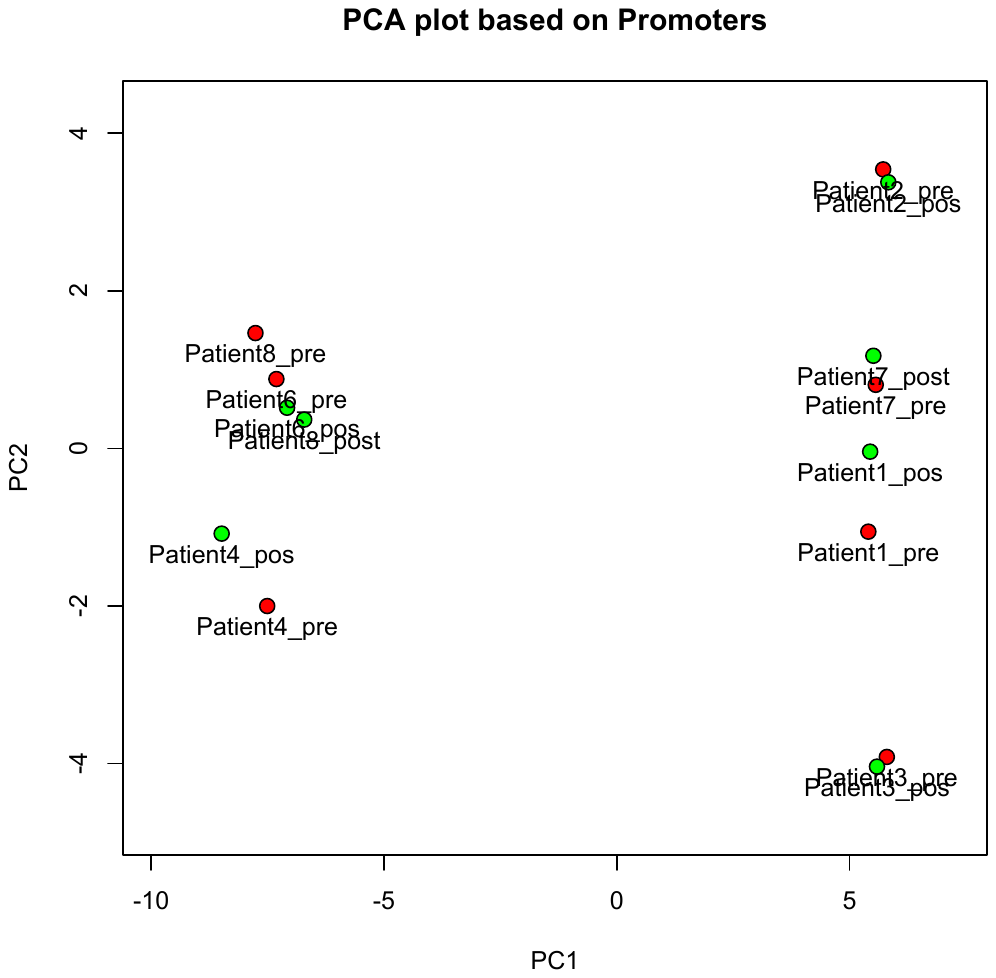

**Supplementary Figure S1.**

Genome-wide PCA of cfDNA methylation profiles (promoter regions only) using (A) all pre- and post-treatment plasma samples; (B) all patients but Patient 5.
